# The sensitivity of SARS-CoV-2 antigen tests in the view of large-scale testing

**DOI:** 10.1101/2020.11.23.20237198

**Authors:** Pavel Drevinek, Jakub Hurych, Zdenek Kepka, Ales Briksi, Michal Kulich, Miroslav Zajac, Petr Hubacek

**Affiliations:** Department of Medical Microbiology, 2nd Faculty of Medicine, Charles University and Motol University Hospital, Prague, Czech Republic; Deptartment of Probability and Statistics, Faculty of Mathematics and Physics, Charles University, Prague, Czech Republic

## Abstract

**Objectives:** Antigen tests have recently emerged as an interesting alternative to SARS-CoV-2 diagnostic PCR, thought to be valuable especially for the screening of bigger communities. To check appropriateness of the antigen based testing, we determined sensitivity of two point-of-care antigen tests when applied to a cohort of COVID-19 symptomatic, COVID-19 asymptomatic and healthy persons.

**Methods:** We examined nasopharyngeal swabs with antigen test 1 (Panbio Covid-19 Ag Rapid Test, Abbott) and antigen test 2 (Standard F Covid-19 Ag FIA, SD Biosensor). An additional nasopharyngeal and oropharyngeal swab of the same individual was checked with PCR (Allplex SARS-nCoV-2, Seegene). Within a 4-day period in October 2020, we collected specimens from 591 subjects. Of them, 290 had COVID-19 associated symptoms.

**Results:** While PCR positivity was detected in 223 cases, antigen test 1 and antigen test 2 were found positive in 148 (sensitivity 0.664, 95% CI 0.599 - 0.722) and 141 (sensitivity 0.623, 95% CI 0.558 - 0.684) patients, respectively. When only symptomatic patients were analysed, sensitivity increased to 0.738 (95% CI 0.667 - 0.799) for the antigen test 1 and to 0.685 (95% CI 0.611 - 0.750) for the antigen test 2. The substantial drop in sensitivity to 12.9% (95% CI 0.067 - 0.234) was observed for samples with the PCR threshold cycle above > 30.

**Conclusions:** Low sensitivity of antigen tests leads to the considerable risk of false negativity. It is advisable to implement repeated testing with high enough frequency if the antigen test is used as a frontline screening tool.

## Introduction

Early detection of SARS-CoV-2 infected individuals is a key factor for making the containment measures effective. Since the beginning of the pandemic, a nucleic acid detection by PCR has become a gold standard of a novel coronavirus disease (COVID-19) diagnostics (1). However, not fast enough turnaround time, a need for laboratory equipment and the shortage of reagents and plastic consumables raised concerns over the time about the PCR as the only frontline tool for testing, especially in surveillance regimes where great numbers of people need to be tested in relatively short time frames (2).

To overcome technical barriers associated with the use of PCR, point of care tests have been considered to complement diagnostic PCR tests and to be used for fast and onsite examination in various settings with suspected outbreaks of COVID-19. These include not only institutions and semi-closed communities such as schools and care homes (WHO interim guidance (3)), but even whole districts and countries, screened in the way of population-wide testing (ECDC Report (4)).

An attractive candidate which meets the logistic criteria for mass testing is an antigen-based detection of SARS-CoV-2 on the principle of immunochromatography. The test is inexpensive, rapid, ready and easy to use. Nevertheless, rapid antigen tests have potential limits in terms of low sensitivity that had been repeatedly documented for other respiratory viruses (5). First reports on the performance of SARS-CoV-2 antigen detection indicated similar findings (6-8), although the manufacturers of commercially available SARS-CoV-2 antigen tests commonly claim sensitivities over 90%. However, these values reflect results of studies done on individuals who meet the criteria for the intended use of the kits, i.e. diagnostics of COVID-19 in patients with clinical symptoms, not in a mixed population of symptomatic, asymptomatic and healthy persons.

In our pilot study, we aimed to evaluate the performance of two antigen tests in a scenario close to the population-wide testing. In this regard, we tested a group of nearly 600 people who had no common epidemiological link between each other.

## Methods

### Subjects

Within a 4-day period in October 2020, we tested 591 individuals of 10 years of age or older, who attended a single collection site, dedicated to the SARS-CoV-2 specimen collection at the Motol University Hospital, Prague, Czech Republic, and consented to the study. The main reasons for their SARS-CoV-2 collection site visit were either the suspicion of COVID-19 infection (273 patients) or contact tracing (290 cases). While 511 persons were referred by general practitioner or public health officer, 54 individuals were self-payers. The mean age of the cohort was 40 years (age range 12 to 78 years), 44.7% were males. Nearly one half of the population (290 subjects) self-reported presence of one or more of the following symptoms: cough, pain of muscles and/or joints, chills, diarrhoea and/or vomiting, elevated body temperature, loss of smell and/or taste. The study was approved by the hospital Ethics Committee (ref no EK-1286/20).

### Antigen tests and PCR

Upon the subject’s consent, we sampled three separate nasopharyngeal swabs and one additional swab from the oropharynx. Two nasopharyngeal samples were used onsite for two antigen detection assays according to the manufacturers’ instructions: Panbio Covid-19 Ag Rapid Test (Abbott, Germany; hereafter referred to as “Ag test 1”) and Standard F Covid-19 Ag FIA (SD Biosensor, Republic of Korea; hereafter referred to as “Ag test 2”). Briefly, the swab was first inserted into an extraction buffer provided with the kit, then the amount of 5 (Ag test 1) or 4 drops (Ag test 2) was loaded on the test device. The results were read after 15 minutes incubation at room temperature by a naked eye (Ag test 1) or after 30 minutes incubation at room temperature on the bench (Ag test 2) in the Standard F200 Analyser (‘read-only’ mode). In order not to unnecessarily lose the sensitivity of the assays, we ran antigen tests immediately upon collecting the sample and without the optional step of inserting the swab into the viral transport medium that may lead in undesirable antigen dilution.

The remaining nasopharyngeal swab along with the oropharyngeal swab were sampled in accordance with the international specimen collection guidelines (CDC, (9)). They were both inserted into the viral transport medium (10) and transported to the hospital microbiology laboratory for the PCR analysis. RNA extraction was performed with Viral Nucleic Acid Extraction kit (Zybio, China) on the EXM3000 instrument (Zybio, China). The extracts were subjected to the reverse transcription PCR, targeting N, E and RdRP/S genes (Allplex SARS-nCoV-2; Seegene, Republic of Korea), run on the CFX96 PCR cycler (Bio-Rad, USA). The sample was deemed positive if at least one of the genes was detected with a threshold cycle (Ct) value ≤ 40; to define a single Ct for a respective sample, we used the lowest Ct out of the three detected targets.

## Results

The PCR positivity was detected in 223 cases (37.7%). Out of them, 168 people had one or more COVID-19 related symptoms (57.9% of all individuals with symptoms and 75.3% of all PCR positive cases), while 55 PCR positive subjects reported no symptoms at the time of sampling. Ag test 1 and Ag test 2 were found positive in 148 and 141 cases, respectively (Table 1).

**Table 1.** Number of positive and negative results when Ag test 1 (Table 1a) and Ag test 2 (Table 1b) are compared to PCR results. Sensitivity and specificity values are reported with 95% CI (in brackets).

|  |  | PCR |  |  |
| --- | --- | --- | --- | --- |
|  |  | positive | negative | Total |
| Ag test 1 | positive | 148 | 0 | 148 |
|  | negative | 75 | 368 | 443 |
|  | Total | 223 | 368 | 591 |
|  |  | Sensitivity 0.664<br>(0.599 - 0.722) | Specificity 1.000<br>(0.990 - 1.000) |  |

|  |  | PCR |  |  |
| --- | --- | --- | --- | --- |
|  |  | positive | negative | Total |
| Ag test 2 | positive | 139 | 2 | 141 |
|  | negative | 84 | 366 | 450 |
|  | Total | 223 | 368 | 591 |
|  |  | Sensitivity 0.623<br>(0.558 - 0.684) | Specificity 0.995<br>(0.980 - 0.999) |  |

Test sensitivity increased to 0.738 (95% CI 0.667 - 0.799) for the Ag test 1 and to 0.685 (95% CI 0.611 - 0.750) for the Ag test 2 if only a subgroup of symptomatic patients (290 subjects) was analysed (data not shown). On the contrary, a low sensitivity value of 0.436 (95% CI 0.314 - 0.567) for either of the Ag tests was found in asymptomatic persons (301 subjects).

The likelihood of detecting the SARS-CoV-2 antigen in a PCR positive person increased with decreasing PCR threshold cycle (Ct) (Table 2). The majority of PCR positive findings (161 of 223, i.e., 72%) had low Ct cycles < 30. In vast majority of cases, these PCR results belonged to patients with COVID-19 associated symptoms (130 of 161 patients). Nevertheless, PCR results with Ct > 30 also comprised mostly the symptomatic patients (38 of 62 cases). Sensitivity of Ag tests was found greater than 80% only for samples with Ct < 30; a substantial drop in sensitivity to mere 12.9% was observed for samples with Ct > 30.

**Table 2.** Sensitivity of Ag test 1 and Ag test 2 in relation to the Ct if samples with higher Ct are added to the samples with lower Ct (Table 2a), and if samples with lower Ct are taken away from the samples with higher Ct (Table 2b).

| PCR Ct | No pts | No symptomatic<br>pts | No asymptomatic<br>pts | Ag test 1<br>sensitivity | Ag test 2<br>sensitivity |
| --- | --- | --- | --- | --- | --- |
| < 20 | 51 | 43 | 8 | 0.922<br>(0.815-0.969) | 0.922<br>(0.815-0.969) |
| < 25 | 121 | 99 | 22 | 0.926<br>(0.865-0.960) | 0.901<br>(0.835-0.942) |
| < 30 | 161 | 130 | 31 | 0.870<br>(0.809-0.913) | 0.814<br>(0.747-0.866) |
| < 35 | 190 | 154 | 36 | 0.779<br>(0.715-0.832) | 0.726<br>(0.659-0.785) |
| ≤ 40 | 223 | 168 | 55 | see Table 1a | see Table 1b |

**Table 2.** Sensitivity of Ag test 1 and Ag test 2 in relation to the Ct if samples with higher Ct 5 are added to the samples with lower Ct (Table 2a), and if samples with lower Ct are taken 6 away from the samples with higher Ct (Table 2b).
| PCR Ct | No pts | No symptomatic<br>pts | No asymptomatic<br>pts | Ag test 1<br>sensitivity | Ag test 2<br>sensitivity |
| --- | --- | --- | --- | --- | --- |
| > 20 | 172 | 125 | 47 | 0.587<br>(0.512-0.658) | 0.535<br>(0.460-0.608) |
| > 25 | 102 | 69 | 33 | 0.353<br>(0.267-0.450) | 0.294<br>(0.214-0.389) |
| > 30 | 62 | 38 | 24 | 0.129<br>(0.067-0.234) | 0.129<br>(0.067-0.234) |
| > 35 | 33 | 14 | 19 | 0.0<br>(0.0-0.104) | 0.03<br>(0.005-0.153) |

## Discussion

With the surge of the SARS-CoV-2 epidemic wave in autumn 2020, novel testing strategies to tackle the community transmission are being sought, including the option of population-wide screening with the aid of antigen tests (4). In our study, we mimicked a situation of mass screening in that we tested each individual, attending the hospital COVID-19 collection site, regardless of the presence or absence of clinical symptoms. Within 4 days, we enrolled nearly 600 individuals out of 800 eligible people who were 10 years or older. Our PCR positivity rate was almost 38% which was well in accordance with over 30% observed on a national level at the time of the study performance (daily reports on (11)).

In our study, we worked with two different antigen tests out of which one enabled europium fluorescence-based detection (Ag test 2), the detection that was believed to improve sensitivity. However, this mode of result visualisation did not have any impact on the change of sensitivity. Overall mean sensitivity values were 66.7% for Ag test 1 and 62.6% for Ag test 2. If the parameter of the presence of clinical symptoms is a criterion for performing the test, sensitivity increased only modestly to 73.8% for Ag test 1 and 68.5% for Ag test 2, while it dropped below 50% if asymptomatic, but PCR positive persons are tested.

Our findings fit well with the known sensitivity characteristics of antigen tests for other respiratory viruses like influenza or respiratory syncytial virus where rapid immunochromatography tests reach the sensitivity of 54.4% and 80%, respectively (12, 13). Such discrepancies between sensitivity values reported in research articles and manufacturers’ leaflets come from differences in selection of tested samples. For instance, a clinical evaluation of the Ag test 2 with the claimed sensitivity of 100% was performed on a positive spiked material, not on real subjects. Interim data on Ag test 1 showed the sensitivity of 85.5% (95% CI 78.2, 90.6), based on testing of 535 patients with suspicion of COVID-19 (14). Of them, 77% were checked by the antigen test within three days from the onset of the symptoms, and 75% had their Ct < 25.

Similarly to Scohy et al. (8), we found out that the main attribute affecting the antigen test sensitivity is a viral load as estimated by the Ct. Lower viral load, represented in our study by high Ct values above 30, became hardly detectable by any of the two antigen tests used. Thus, patients with samples of late Ct values would be largely left undiagnosed, although many of them also presented with clinical symptoms in our study. Regardless of their clinical state, it is important to point out that they all might be also infectious as documented in studies on asymptomatic and presymptomatic persons (15, 16).

Because of the recent findings on infectivity (17), we can speculate that patients with low Ct were actually at the end of their infection stage and no longer posed the risk to others. However, our patient records show that only 5 of 62 patients with Ct > 30 visited the collection site to monitor the course of their infection (3 of them had still clinical symptoms), so we can assume that most of them were near the onset of the disease and cannot rule out their infectivity. In reference to the aim of our study, i.e. to check the effectiveness of the mass testing with antigen tests, another limitation of the study is the bias in selection of the subjects who were in most cases indicated for the examination due to symptoms or contact tracing. One can expect that the population-wide screening would include higher rate of asymptomatic people with low viral loads as well as healthy persons which in turn would lead in even higher false negativity and false positivity rates (18).

To conclude, in our opinion, the risk and rate of false negativity of antigen tests may have a significant negative impact on the effectiveness of outbreaks containment as it is crucial to early identify any positive person, including the ones with initially low viral load. Not to miss them, a single round of testing, which is likely the case when a population-wide screening is ordered, seems insufficient and inadequate. Instead, the strategy based on repeated testing with high enough frequency (2) needs to be implemented if the antigen test is used as a frontline screening tool.

## Data Availability

All data are included in the manuscript.

## Transparency declaration

Supported by the Ministry of Health of the Czech Republic - conceptual development of research organization Motol University Hospital, FNM. All authors report no conflicts of interest relevant to this article.

## Acknowledgements

The authors are grateful to I. Adam, M. Sadilkova, K. Smolkova, M. Kalantay, I. Prosova, S. Strba, D. Kovarikova, B. Dzerengova, A. Hutnan, A. Shaker, V. Casarova and other staff members of Motol collection site for excellent technical assistance.

